# The effect of COVID-19 lockdowns on fertility in the Democratic Republic of the Congo

**DOI:** 10.1101/2022.12.16.22283557

**Authors:** Shuo Feng, Gabriel Kyomba, Serge Manitu Mayaka, Karen Ann Grépin

## Abstract

Most countries implemented public health measures, including lockdowns, during the COVID-19 pandemic. It has been speculated that the pandemic will affect fertility, but the direction, magnitude, and mechanisms of these effects are not well understood. Using data from the national health management information system and an augmented synthetic control methodology, we examined the impact of a lockdown of Kinshasa in April 2020 on the subsequent fertility of women, which we proxy by the number of births in health facilities months after the policy was implemented. Seven months after the lockdown, we see a large increase in births in Kinshasa, as compared to control areas, which at its peak represents an additional 5000 monthly births, or a 45% increase relative to baseline. We also observe increases in complimentary maternal health services but not in other health services. Increased births were observed among women both older and younger than 20. Lockdown policies have likely affected fertility and future pandemic preparedness plans should anticipate the effects find strategies to mitigate any negative unintended effects.

## 1. Introduction

The COVID-19 pandemic led many countries, including many low- and middle-income countries (LMICs), to impose far-reaching public health measures in hopes of reducing viral transmission. However, bans on gatherings, school and business closures, and widespread mobility restrictions also had unprecedented economic and social effects. A large proportion of households in LMICs experienced declines in employment and income (Bundervoet et al., 2022; Egger et al., 2021). About a third of children in households in 31 LMICs surveyed at the end of 2020 were still not learning due to school closures (Bundervoet et al., 2022). Lockdowns, or other stay-at-home orders, were particularly consequential, especially in Sub- Saharan African countries (IMF, 2020).

Beyond the economic and social effects, government response measures also likely affected women’s sexual and reproductive outcomes in these settings, including changes in fertility intention, unintended pregnancies, disruptions in the use of sexual and reproductive health services, and worsened maternal and child health outcomes. Studies of fertility patterns during and in the aftermath of the 1918 influenza pandemic generally found declines in fertility, and while early evidence of the impact of the COVID-19 pandemic on fertility rates on high-income countries (HICs) also generally found lower fertility rates in early pandemic period, substantial heterogeneity was also observed (Aassve et al., 2021). As was the case a century ago, less is known about the impact of the pandemic on fertility in very low-income settings where vital registration systems are incomplete. Survey evidence from Kenya did not find any evidence that the pandemic had led to important changes in fertility intentions (Zimmerman et al., 2022).

In many LMICs, there are other worrying signs that sexual and reproductive health outcomes have been affected. A systematic review has demonstrated that maternal and perinatal health outcomes worsened in the months immediately following the start of the pandemic across both high and LMIC settings (Chmielewska et al., 2021). A lockdown implemented in Nepal in March 2020, led to steep declines in institutional delivery rates, as well as increases in rates of institutional stillbirth and neonatal mortality (Kc et al., 2020). However, the specific mechanisms through which the COVID-19 pandemic, as well as government response measures, may have affected sexual and reproductive outcomes are less well understood. Prior to COVID-19, studies of the impact of infectious disease outbreaks on sexual and reproductive health outcomes had identified the negative impact of school closures on teenage pregnancies and the short-term disruptive effect of outbreaks on the use of sexual and reproductive health services (Bandiera et al., 2020; Elston et al., 2017; Wilhelm and Helleringer, 2019).

Following their successful application in Wuhan, China, many countries adopted mobility restrictions during the early waves of the pandemic. The rationale for the use of so-called “lockdown” measures largely stems from the fact that the SARS-CoV2 virus spreads easily from asymptomatic patients and it has a relatively long incubation period. Several countries, including South Africa and India, implemented national level lockdown policies. Despite the widespread use of measures, little is known about how such measures may have affected sexual and reproductive health outcomes. There is very strong evidence that lockdowns were associated with increases in intimate partner violence (IPV). In India, areas of the country with the highest levels of mobility restrictions recorded large increases in domestic violence (Ravindran and Shah, 2020), which could have also affected rates of unintended pregnancies.

Although there is no commonly agreed definition of a “lockdown”, the term has been frequently used during the COVID-19 pandemic to refer to a range of mobility restrictions, including stay-at-home orders and curfews but the term has also been used to describe other forms of geographic confinement (e.g. cordon sanitaires), forced home confinement, as well as work-from-home policies. The use of such measures was widespread during the pandemic, including in Sub-Saharan African (SSA) countries (Haider et al., 2020).

Like many SSA countries, the Democratic Republic of the Congo (DRC) has been greatly affected by the pandemic, however, the health effects have been less well documented. It is clear, however, that the country has had important social and economic effects due to the measures. As of April 27, 2022, the DRC had officially recorded over 86,000 cases of COVID-19 and over 1,300 deaths^1^, however, it is widely believed that this is only a fraction of the true number of cases. Testing was not widely available and was only available in a small number of cities (Ditekemena et al., 2021b). Testing rates were very low by international standards (Juma et al., 2020). As such, the true number of cases is likely orders of magnitude larger than the officially confirmed number of cases. Indeed, seroprevalence data suggests that the virus had been more prevalent in the country during the first wave than would be suggested based on official case counts (Nkuba et al., 2021). Vaccination rates, as of April 2022, were amongst some of the lowest rates in the world.

Against this backdrop, this study evaluates the effects of lockdown on the fertility of women in the DRC, which we proxy using the number of births observed in health facilities many months after the implementation of the measure. Specifically, we compare the subsequent births of women in Kinshasa, where the most stringent lockdown measures had been introduced in April 2020, to those of women in other parts of the country using an augmented synthetic control methodology. Although we might expect the use of health services to also be directly affected by the lockdown, if the lockdown were to directly affect fertility, we would only expect to see the effects of the lockdown many months after it was implemented. As school closures had been implemented at the national level, it is important to note that all schools had been closed in the DRC at the time of the lockdown in Kinshasa, therefore any effects that we observe must be interpreted as the impact of a lockdown in a setting in which schools were closed. However, this is not an estimate of the impact of school closure itself. Below we first describe our data and methods, present our main findings, and then discuss and conclude.

## 2. Data and Methods

### 2.1 Study context

The DRC is one of the largest and most populous countries in Sub-Saharan Africa (SSA), and it is also one of the poorest. Of the roughly 90 million people who live in the country, at least 17 million (19%) of them live in Kinshasa, the capital city, although the exact number is not known. It has faced a long history of armed conflict, especially in the eastern part of the country. Outbreaks of infectious diseases are common in the DRC: when the COVID-19 pandemic hit in early 2020, the DRC was already dealing with a large-scale outbreak of Ebola Virus Disease in the Eastern part of the country. These experiences had helped it to prepare for the pandemic, but its weak health system and other public infrastructure limited its ability to effectively respond (Ditekemena et al., 2020; Mobula et al., 2020).

Fertility remains high in the DRC, and it is one of the few countries in the world that has not begun its demographic transition (Romaniuk, 2011). In fact, the total fertility rate increased between 2007 and 2013-14 to 6.6 children per woman (Kwete et al., 2018). As a result, population growth rates remain high. Although rates of uptake of family planning have increased and are higher in Kinshasa, the modern contraceptive prevalence rate overall in the country remains low (Kwete et al., 2018). However, according to the most recent Demographic and Health Survey (DHS) in the DRC, which was conducted in 2013-14, rates of institutional delivery are relatively high by regional standards – almost 80% of women nationally gave birth in a health facility (Gebremichael and Fenta, 2021; Modernité et al., 2014). In Kinshasa, nearly 98% of women had given birth in a health facility.

The DRC confirmed its first case of COVID-19 on March 10, 2020, in an adult male who had recently returned to Kinshasa from France (Nachega et al., 2020). The government immediately began to implement measures to contain the virus, first by implementing health screening at the airport. National-level school closures were announced on March 18, along with additional measures such as the closing of restaurants and bars, bans on large gatherings, and limitations on travel in and out of Kinshasa. On the following day, all travel in and out of Kinshasa was banned. As the initial measures were not seen as sufficient and following reports of a COVID-19 death and the detection of additional cases, the national government declared a state of emergency on March 24, 2020. On the following day, all international borders were closed and domestic flights between Kinshasa and the rest of the country were suspended. The nationwide state of emergency declaration stayed in force until July 22, 2020, when a reopening plan was also announced. Nationally, schools only reopened on August 10 after approximately 5 months of closure.

From the start of the pandemic, Kinshasa has been seen as the epicenter for COVID-19 in the country. It was also the site of the strictest lockdown imposed during the pandemic. The Governor of Kinshasa initially announced a month-long lockdown of the entire capital region starting on March 28, but the measure was called off due to concerns about the practicality of the strategy. Gombe, a relatively central and affluent neighborhood that was seen as the local hotspot of the outbreak, was subject to a strict lockdown that started on April 6 (Nachega et al., 2020). During the lockdown, all businesses were closed, and public transportation was suspended. Hospitals and pharmacies, however, remained open. Everyone was required to stay at home all day, except for essential outings, such as seeking medical care. Gombe is also the location of the Kinshasa Provincial Hospital, a high-volume referral facility that services the entire Kinshasa Province (Hategeka et al., 2021). The lockdown was initially only supposed to last 2 weeks, but on April 22, the measures were only slightly loosened, and they were not fully lifted for another 6 weeks when businesses and restaurants reopened in Gombe on June 28, 2020.

Lockdowns, curfews, and other mobility restrictions were also used in other parts of the DRC, but they were not as stringent, nor were they as long, as those put in place in Kinshasa. Lubumbashi, the third-largest city in the DRC and the capital city of Haut-Katanga Province, was the second place in the DRC to detect a case of COVID-19 on March 21, 2020. Two days later, on March 23, Lubumbashi was placed in a 48-hour lockdown after two suspected cases of COVID-19 had disembarked from a flight from Kinshasa. On April 20, Lubumbashi and Lualaba, another urban center in Haut-Katanga Province, both imposed a curfew that prevented people from leaving their homes from 10 p.m. to 5 a.m. On April 29, Lubumbashi and Kasumbalesa, another Haut-Katangan town, were placed on a 24-lockdown to facilitate contact tracing of COVID-19 cases. A 3-day lockdown was also instituted in Lubumbashi, Kasumbalesa, Likasi, and Kipushi on July 11, 2020.

Cases had also been identified earlier in the Eastern part of the country, especially in North Kivu Province, the centre of an ongoing outbreak of Ebola at the time. On May 20, the Provincial Government of North Kivu placed a nightly curfew on the city of Goma, and travel in and out of the city was suspended. People were also encouraged to stay at home as much as possible and there was a ban on mass gatherings, but businesses remained open and public transportation continued to run. To our knowledge, no other areas in the DRC were subject to formal lockdown measures during the initial wave of the pandemic in early 2020.

### 2.2 Data

To estimate the impact of the lockdowns, we used data on the total number of births and other health services delivered in health facilities, which we extracted from the DRC’s Health Management Information System (HMIS) – a routine health information system based on the open-sourced District Health Information System 2 (DHIS2) platform. While the true number of births amongst women in the DRC is not directly observable in the HMIS as we only observe those that take place in a health facility, rates of institutional deliveries are very high in Kinshasa at 98% (Ministère du Plan et Suivi de la Mise en œuvre de la Révolution de la Modernité et al., 2014). A previous study conducted in Kinshasa that looked at the immediate impact of the pandemic on the use of health services did not find any effect of the lockdowns on institutional delivery rates in the months following the lockdown (Hategeka et al., 2021). Other health services, however, had been adversely affected in the short run.

From the HMIS, we extracted monthly data on the number of deliveries, including the total number of deliveries for the entire population as well the number of deliveries to mothers under the age of 20, the number of fourth antenatal care visits (ANC), which are typically scheduled and usually take place around 36-38 weeks of gestation, and the number of postnatal care visits delivered within 6 hours of birth (PNC). We also extracted the total number of doses of the *Bacillus Calmette-Guérin* (BCG) vaccine, which is used to protect against tuberculosis and is typically given at birth in the DRC, and the *Diphtheria-Pertussis-Tetanus-Polio-Hepatitis B-Haemophilus Influenzae Type B* (DTP-HepB-Hib) vaccine, which is usually given in the first few weeks of life. We selected these vaccines because children born in health facilities should be administered these vaccines at birth or shortly thereafter during follow-up health visits. Finally, to control for changes in the delivery of other services, we also collected data on other health services, including total outpatient visits, which excludes maternal health service visits, and visits for diarrhea. While these types of visits may have been affected by the pandemic, we would expect disruptions to these types of health services to occur during or in the immediate aftermath of lockdowns, not with a delay that is suggestive of a human gestational period. We collected data from January 2018, or a little more than 2 years before the start of the pandemic, up until February 2022. More details on how the data used in this study are collected and aggregated are available elsewhere (Hategeka et al., 2021; Yuen W Hung et al., 2020).

Births are recorded in the HMIS as follows. In each health facility, register books typically collect information on all births recorded in the health facility. The total number of births recorded in a month is then manually calculated and recorded onto tally sheets that are then sent to the health zone office, where the data are inputted into the HMIS. Limited information on the mothers is captured in this system, only the facility of birth, and whether the mother was younger than or older than 20 years old. We are therefore limited in our ability to address questions about the specific groups most affected by the lockdown.

Although there have been significant improvements in data quality in the DRC HMIS in recent years, data from most national RHIS in LMIC settings face common challenges, including outliers and missing data (Yuen W. Hung et al., 2020). To prepare our data for analysis, extreme values were first detected and removed based on a random forest model trained using the baseline data, and then we used a seasonally decomposed imputation algorithm to fill in the missing values (Feng et al., 2021). To obtain official data on the dates of policy implementation of the lockdown, we used government websites, news media, published reports, and academic journals. Our local authors also confirmed these dates with their colleagues.

Our dataset is thus the number of births and other health services that were delivered in each health facility in the DRC per month. Our sample includes mainly public health facilities, which include health centres, health posts, and hospitals that report into the central HMIS system. Most private health facilities do not regularly report into the DHIS2 system, although some private facilities are regularly captured in the system. In addition, facilities that were closed or stopped reporting before the start of the pandemic (4,574 out of 18,329 facilities, or 29.60%) were also excluded from the study. Monthly aggregated data from January 2018 to February 2020 were considered pre-pandemic, or our baseline period, and April 2020 was set as the start of the lockdown in Kinshasa. We consider all data among the rest of the facilities to be the baseline, upon which we perform data cleaning and construct synthetic controls to mimic the trajectory of Kinshasa (discussed in the following section). Haut Katanga and North Kivu were also excluded from the analysis because they were also exposed to shorter lockdowns and curfews and thus could have also been affected by the measure. They were also removed from our baseline sample. We have also estimated our models with them included, and it has little effect on our results.

### 2.3 Statistical Methods

As our goal was to investigate the impact of lockdown on fertility, we defined women who give birth anywhere in Kinshasa to have been exposed to the lockdown, whereas women who gave birth elsewhere were not exposed to the lockdown. Given a standard gestational period is roughly 40 weeks from a women’s last menstrual period, which is generally two weeks before conception, if the lockdowns were to have had an effect, we would expect to start to see an effect approximately 7-8 months later, peaking approximately 9-10 months later. While we do not directly observe actual fertility in the population, we do observe the number of women giving birth in health facilities per month. Assuming no major changes in the proportion of women who give birth in a health facility over time, we interpret the proportional increase in the number of births observed in our dataset as a proxy for the proportional increase in fertility that results from the lockdown. In our limitations section below, we discuss the implications of our findings if we assume that women who are the most likely to be affected by the lockdowns became less likely to give birth in clinics over time.

To estimate the impact of the lockdowns on fertility, we use a synthetic control method (SCM), which is a method that has been developed to estimate the causal effect of interventions on outcomes. In our study, we are interested in the average treatment effect on the treated (ATT), which is the difference between the observed outcomes of the treated unit (i.e., Kinshasa which is exposed to lockdown) and its counterfactual outcomes in the absence of the intervention, which is not observed and thus needs to be estimated via a SCM (Abadie et al., 2015). A traditional difference-in-differences-like approach approximates this causal effect by finding a control unit in the sample and estimates the ATT as the difference in the outcome between the treated and the chosen control unit before and after the intervention. However, this type of method requires us to find a control unit that is very similar to the treated unit in all ways, which is rarely possible because, as in the case of this study, there is no one single province in the control that is comparable to our treated province in every way (e.g., population, geographics, economics, policy, etc.).

Instead of identifying just one single control unit to be compared to the treated, the SCM constructs an artificial or a “synthetic” control of the treated unit using a weighted average of the control areas in the sample that closely mimics the trajectory of the treated unit up to the intervention. If we manage to construct such a synthetic control that can reproduce the outcomes from the treated unit at all time periods before the intervention, we can then attribute any divergence found in the post-intervention outcomes between the treated and its synthetic control to the intervention (Arkhangelsky et al., 2021). A typical SCM requires the control unit-level weights to be non-negative and sum to one to avoid extrapolation. However, under this restriction, we may not always be able to construct a weighted average of the controls that matches the treated units well, and as a result, the estimated treatment effect following the intervention does not make sense (Abadie et al., 2015).

An augmented synthetic control method (ASCM) addresses this limitation of the traditional SCM approach by “augmenting” the original SCM using an outcome model (usually a Ridge-regularized regression model) to estimate and correct the bias due to the poor pre-intervention fit where only non-negative weights are allowed (Ben-Michael et al., 2021). By allowing negative weights, ASCM is usually able to achieve a better match between the weighted average of the controls and the treated unit of interest. Note that here extrapolation resulting from allowing negative weights is not a problem because any potential bias introduced by this extrapolation is penalized by the Ridge regression (Cole et al., 2020).

Formally, let *A ∈* {0,1} denote the treatment assignment, where *A*= 1 refers to the treated province, i.e., Kinshasa, and *A*= 0 for all the other control units without a lockdown. Let *i ∈* {1, …, *N*} denote each province in the DRC with *N* being the total number of provinces, and let *t* ∈ {1, … *t*/, …, *T*} denote each month throughout our study, where t/ = 28 is the month of lockdown in Kinshasa, and we set the last month to be *T*= 44 which is 16 months after the lockdown, or August 2021. Under a general SCM setting, we establish a latent factor model as follows (Athey et al., 2021; Xu, 2017) :

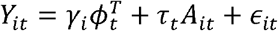

where *τ*_*t*_ is our estimate of interest, or the ATT, that is, the averaged difference between the observed number of deliveries in Kinshasa post-intervention versus the counterfactuals of Kinshasa in the absence of the intervention. In addition, *γ*_*i*_ denotes a vector that contains province-level latent factors (e.g., relative population size, economic and social characteristics of each province), *¢*_*t*_ contains latent time-related factors (e.g., population growth rate, seasonality), and *∈*_*it*_ denotes the idiosyncratic errors that are assumed to have mean 0 conditioning on the treatment assignment *A*_*it*_ and the mixing of latent factors 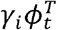.

To estimate the standard errors and construct a 95% confidence interval for inference, we further assume that the errors {*∈*_*it*_ *}* are stationary and weakly dependent. An estimate of standard error and confidence intervals can be then obtained through block permutations under the null *H*_0_: *T*_*t*_ =0 for every *t*= *t*/,…, *T* (Chernozhukov et al., 2021).

### 2.4 Placebo Tests

In addition to the examination of standard errors and confidence intervals, we also wish to evaluate how well the SCM approach applies to our data in general. Abadie et al. (2015) suggest testing the validity of the treatment effect estimates through placebo studies. The logic of these placebo tests is that if we artificially re-assign the intervention to a control province, we should expect a treatment effect that is much smaller in size than the one we obtained using the true treated unit. If, however, the estimated effects obtained by pretending the controls as the treated are of similar or even larger magnitude, then our confidence in the estimated treatment effect from the true treated unit is also greatly undermined. We conducted placebo tests and report the results in the Findings section below.

Using ASCM, we, therefore, estimate the total number of additional births per month relative to the pre-pandemic and pre-lockdown baseline number of births in Kinshasa. In the pre-pandemic period, on average we observed approximately 12,035 total births per month in Kinshasa, of which, 1,947 births (16%) were among women under the age of 20. In addition, we also calculate the percentage increase in births per month, again relative to the pre-pandemic reference period. We repeat these calculations for each of the additional health services also investigated.

## 3. Results

Table 1 summarizes the monthly number of deliveries in our sample. It also reports the estimated weights from the ASCM, as well as the weights that we would have obtained using a regular SCM for comparison.

**Table 1:** Summary statistics of the dataset and ASCM weights

| Province | Average Monthly Deliveries in the Baseline | Weights from ASCM | Weights from Regular SCM |
| --- | --- | --- | --- |
| **Kinshasa (Target Province)** | 12,035 |  |  |
| **Kongo Central** | 7,505 | 1.39 | 0.58 |
| **Sud Kivu** | 18,306 | 0.51 | 0.40 |
| **Maniema** | 7,633 | 0.47 | 0.00 |
| **Kasai** | 16,209 | 0.38 | 0.02 |
| **Haut Uele** | 4,270 | 0.18 | 0.00 |
| **Haut Lomami** | 11,496 | 0.16 | 0.00 |
| **Lomami** | 10,931 | 0.14 | 0.00 |
| **Mongala** | 9,329 | 0.08 | 0.00 |
| Maindombe | 6,012 | 0.01 | 0.00 |
| Bas Uele | 2,685 | -0.01 | 0.00 |
| Lualaba | 9,683 | -0.04 | 0.00 |
| Tshuapa | 6,343 | -0.05 | 0.00 |
| Kwango | 8,028 | -0.05 | 0.00 |
| Sankuru | 5,599 | -0.05 | 0.00 |
| Kasai Central | 16,484 | -0.07 | 0.00 |
| Equateur | 7,068 | -0.13 | 0.00 |
| Tshopo | 5,776 | -0.17 | 0.00 |
| Sud Ubangi | 8,862 | -0.18 | 0.00 |
| Kwilu | 15,808 | -0.21 | 0.00 |
| Nord Ubangi | 4,538 | -0.23 | 0.00 |
| Kasai Oriental | 12,581 | -0.26 | 0.00 |
| Tanganyika | 5,136 | -0.41 | 0.00 |
| Ituri | 15,051 | -0.44 | 0.00 |
| Haut Katanga (Excluded from Analysis) | 17,945 |  |  |
| Nord Kivu (Excluded from Analysis) | 24,050 |  |  |

In Figure 1, we illustrate the estimated monthly average difference in births (ATT) between Kinshasa and its synthetic control before and after the lockdown in Kinshasa (April 2020). We expect the ATT to be small before the intervention but large afterward if there is an effect of the lockdown.

**Figure 1a.**
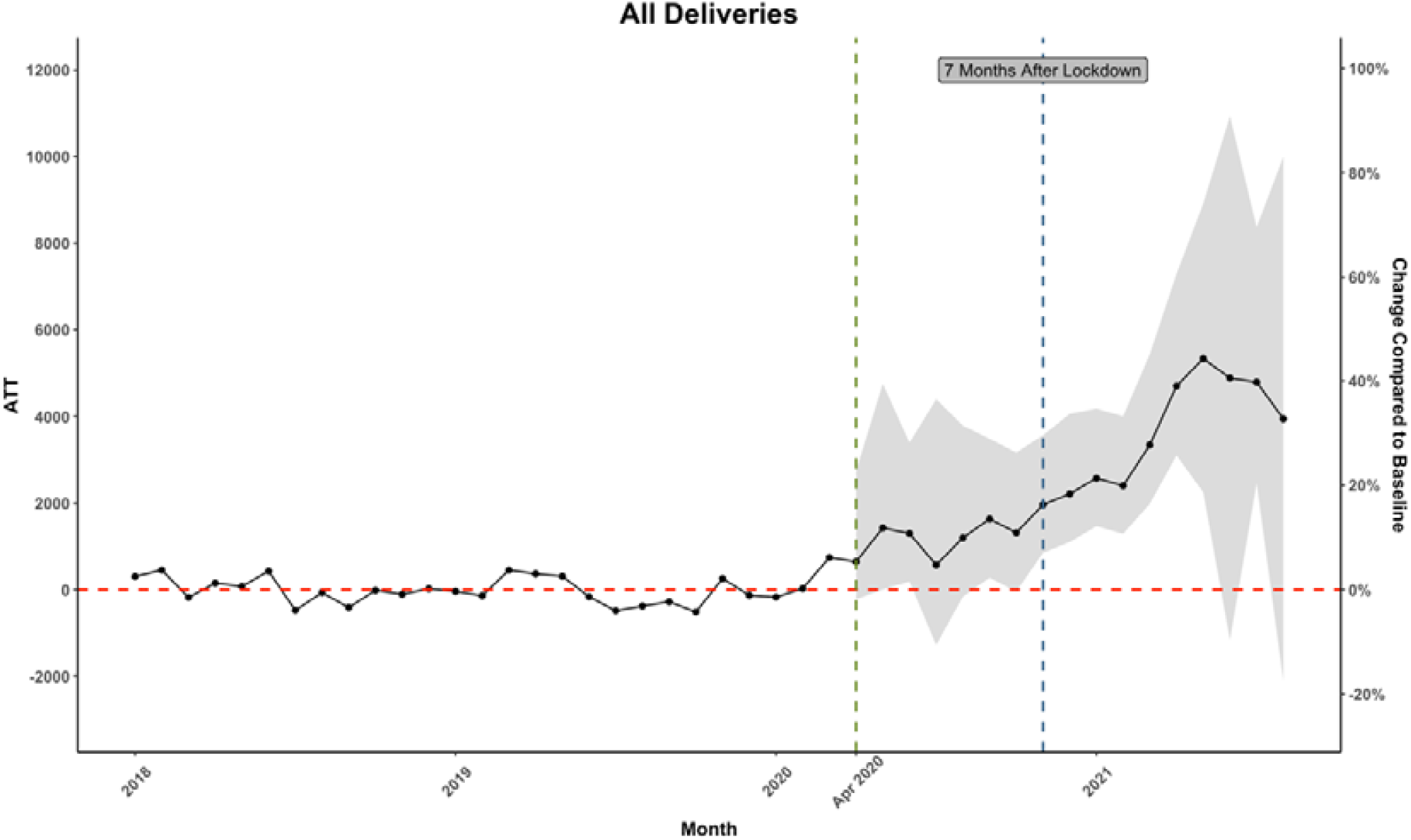
Estimated ATT in All Deliveries in Kinshasa.

**Figure 1b.**
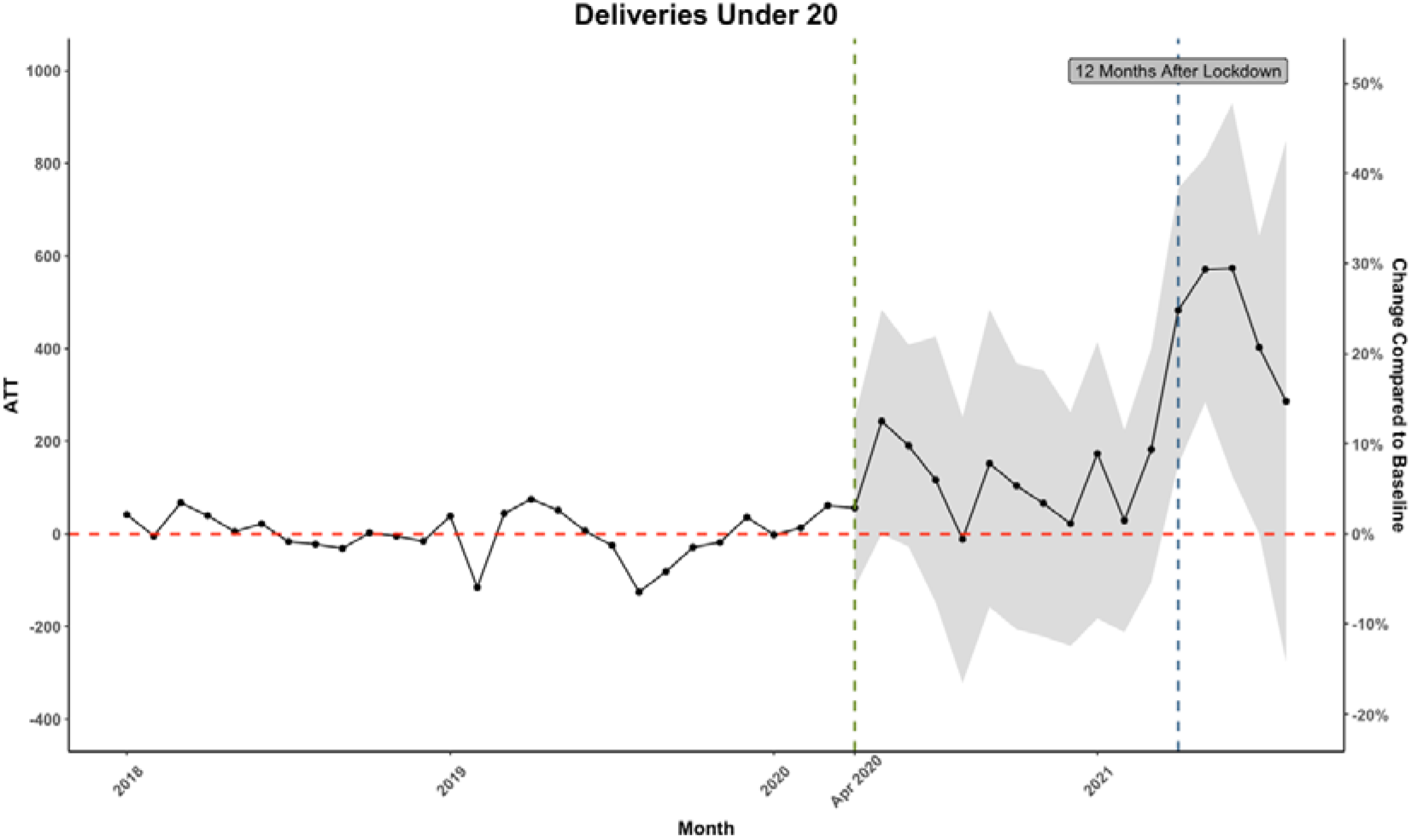
Estimated ATT in Deliveries Under 20 in Kinshasa.

Figure 1a plots the estimated ATT in all deliveries over time in Kinshasa. We observe that our model was able to closely mimic the trajectory of births in health facilities in Kinshasa in the pre-lockdown period. Following the lockdown, we begin to see an increase in the number of births in Kinshasa as compared to its synthetic control, however, we do not see any significant increases until at least 7 months after the lockdown. Births in facilities continue to increase for approximately 5-6 more months, before declining and no longer being statistically significant. At its peak, Kinshasa recorded an excess of about 5,000 additional births per month, from a base of on average 12,035 births per month in the pre-lockdown period, representing an approximately 45% increase in the monthly birth rate.

Figure 1b plots the same ATT estimates, but this time only for mothers under the age of 20. While these data are noisier, we do see a large increase in births, which reaches statistical significance 12 months after the lockdown. Again, this represents a large proportional increase of about 20-30% as compared to the baseline levels.

Figure 2a investigates the rates of ANC services while Figure 2b investigates rates of PNC services. While we do not generally observe any short-term increase in ANC visits, there does appear an increase in the number of ANC4 visits 11 months after the lockdown.

**Figure 2a.**
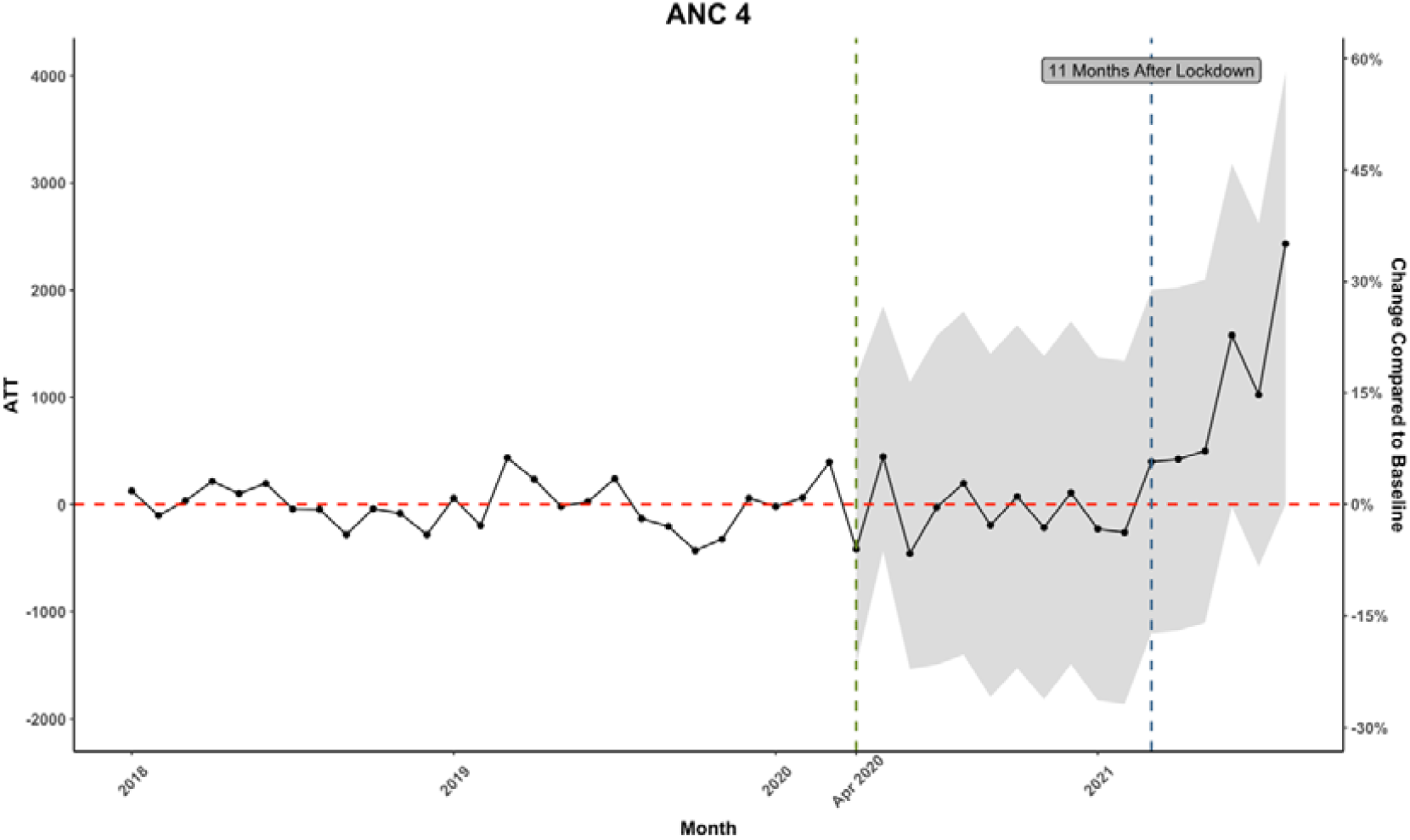
Estimated ATT in ANC4 Visits in Kinshasa.

**Figure 2b.**
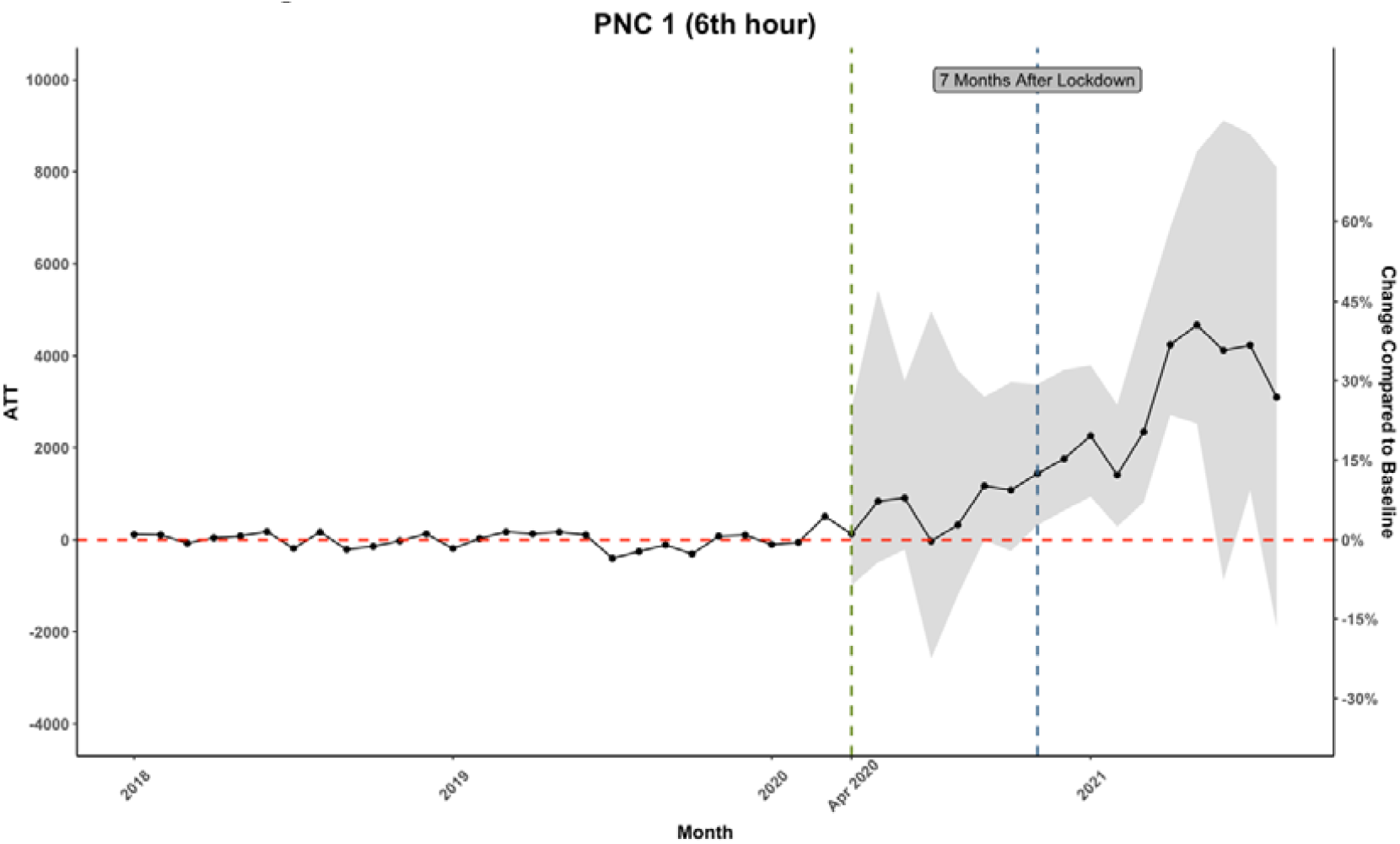
Estimated ATT in PNC1 Visits in Kinshasa.

For PNC, Figure 2b shows the estimated ATT for the first PNC visits that happen within 6 hours of the delivery. We observe a very similar pattern as with overall deliveries, both of which started to increase 7 months after the lockdown. This is consistent with our observed trends in increased facility births as women who are already in a facility are likely to remain to receive their first PNC visit within 6 hours. On the other hand, we do not observe such a strong significant increase in ANC visits partly because they are optional, and women may tend to avoid attending these sessions when the pandemic is still around, especially when the second wave of COVID-19 hit the DRC in December 2020.

Further evidence of an increase in facility-based deliveries is also seen in our data on vaccinations of children. If there were more births, we would expect to see an increase in the number of vaccinations given to these children around the same time. In Figure 3a, we present the estimates of the ATT for BCG vaccinations, a vaccine that is recommended to be given at birth in the DRC. Although there is some evidence of an early decline in the rates of BCG vaccination post-lockdown, after 9 months we see a large and significant increase in vaccinations on the order of approximately 3000 additional vaccines. In Figure 3b, we present the results for the DTP-HepB-Hib1 vaccine, which is given in the weeks immediately following birth, we begin to see a large increase in the number of vaccines administered per month approximately 9 months after the lockdown. We observe about 2500 additional vaccines administered per month relative to the baseline numbers of vaccines.

**Figure 3a.**
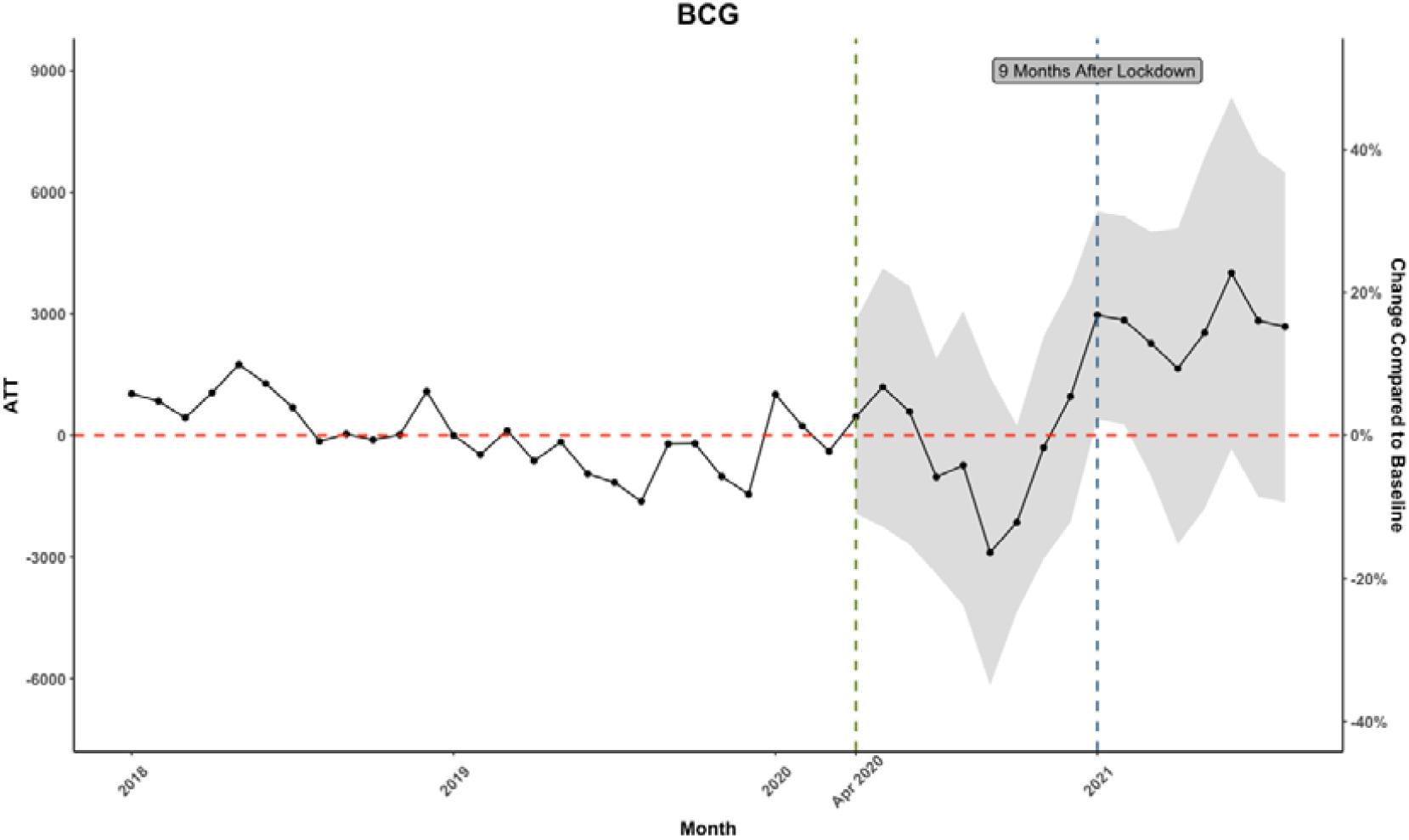
Estimated ATT in BCG Vaccination in Kinshasa.

**Figure 3b.**
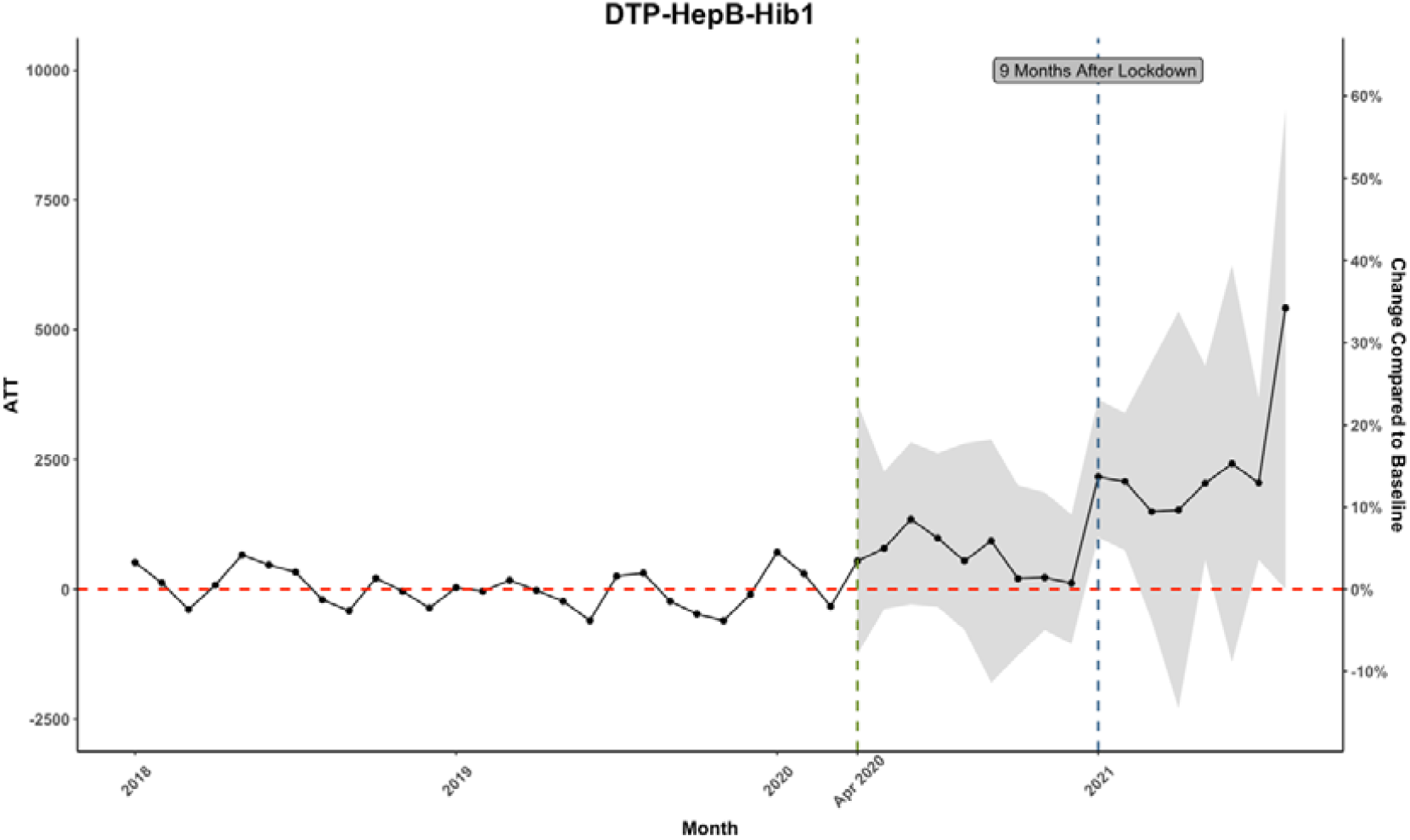
Estimated ATT in DTP-HepB-Hib1 Vaccination in Kinshasa.

Finally, in Figure 4 we investigate other health services that should not have been affected by the lockdown many months after its implementation. In Figure 4a, we investigate the number of cases of diarrhea treated in health facilities, however, we observe no significant increases in the use of this service any time after the lockdown in Kinshasa. In Figure 4b, we investigate total outpatient visits. We observe some increases after 10 months of the lockdown; however, this may be attributable to the other health services that went up along with an increase in deliveries, such as the vaccination and ANC/PNC visits. Regardless, the increases in outpatient visits are not statistically significant at any time after the lockdown.

**Figure 4a.**
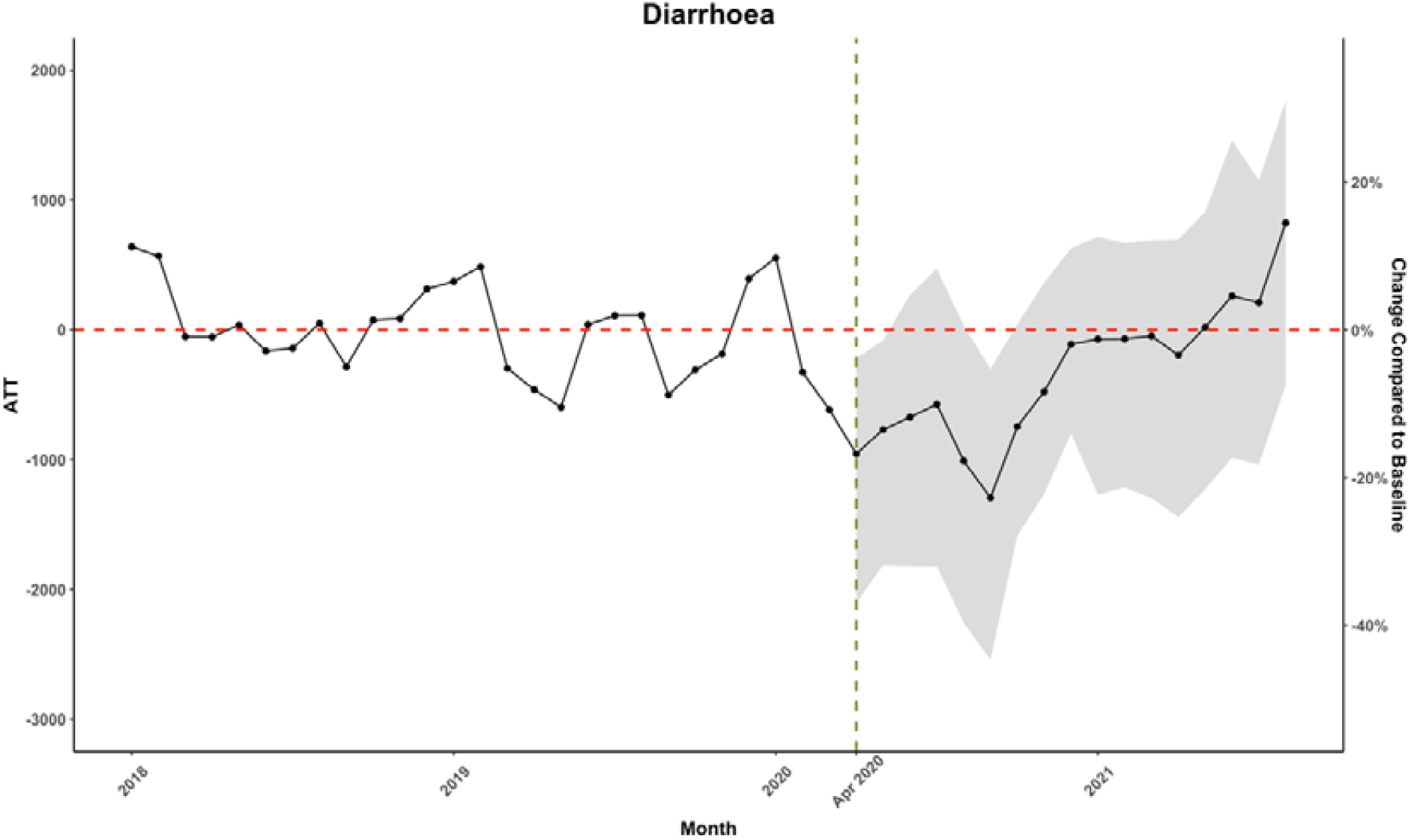
Estimated ATT in Diarrhea Cases in Kinshasa.

**Figure 4b.**
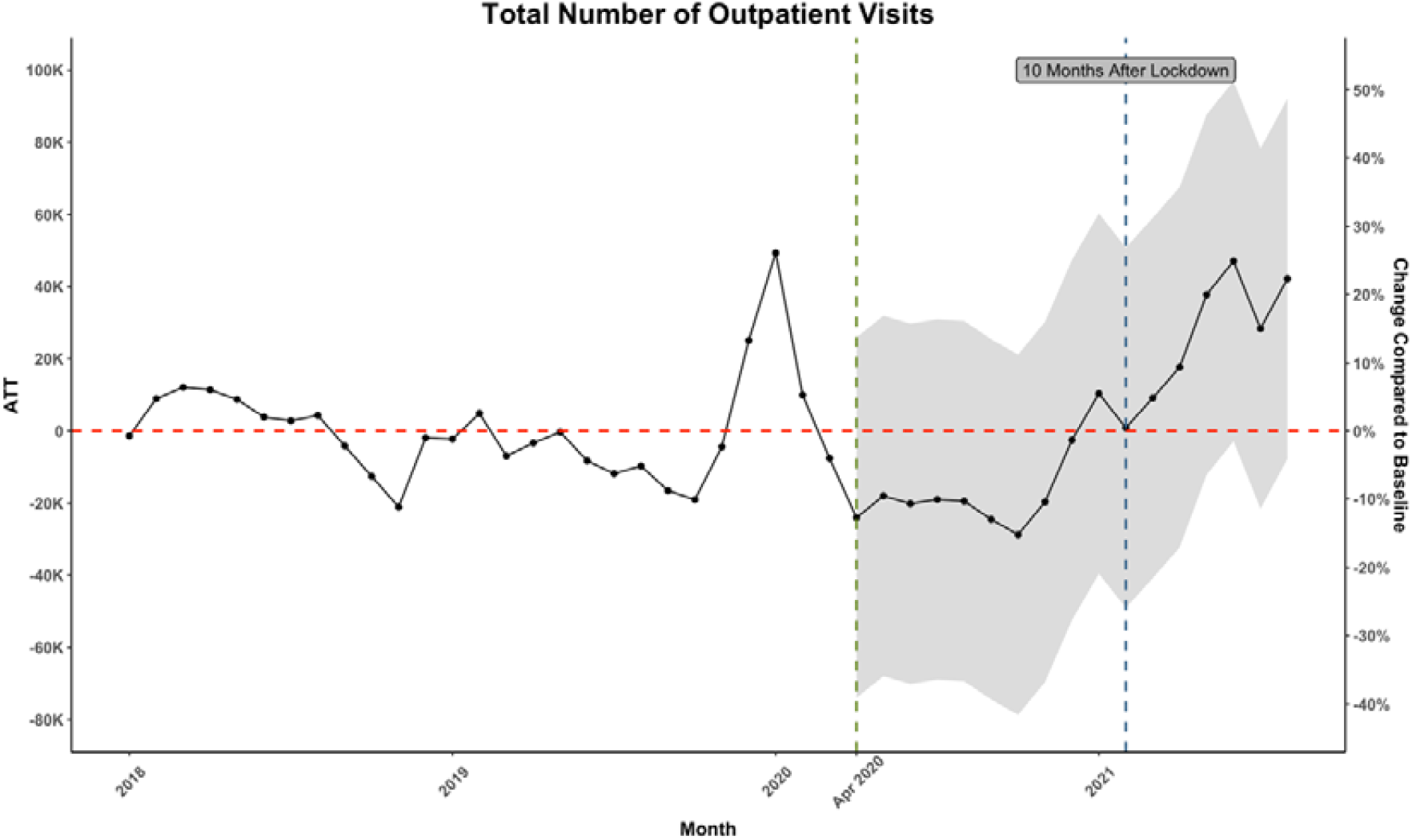
Estimated ATT in Total Number of Outpatient Visits in Kinshasa.

As discussed in the methods section, if there were a true treatment effect in Kinshasa, we would expect to see the treatment effect from Kinshasa to be at the extreme end in the distribution of estimated treatment effects by pretending different provinces in the population as the treated in a placebo test. Figure 5 is generated by bootstrapping the control units and in each iteration, we artificially assign the treatment to one control unit and estimate its treatment effect. Indeed, the estimated ATT in Kinshasa is the largest in size – by far at all periods after the intervention, which provides evidence supporting a true treatment effect resultant from the lockdown in Kinshasa. All pre-intervention estimates of ATT are clustered and fluctuating around 0, which again indicates a good fit of the ASCM model.

**Figure 5.**
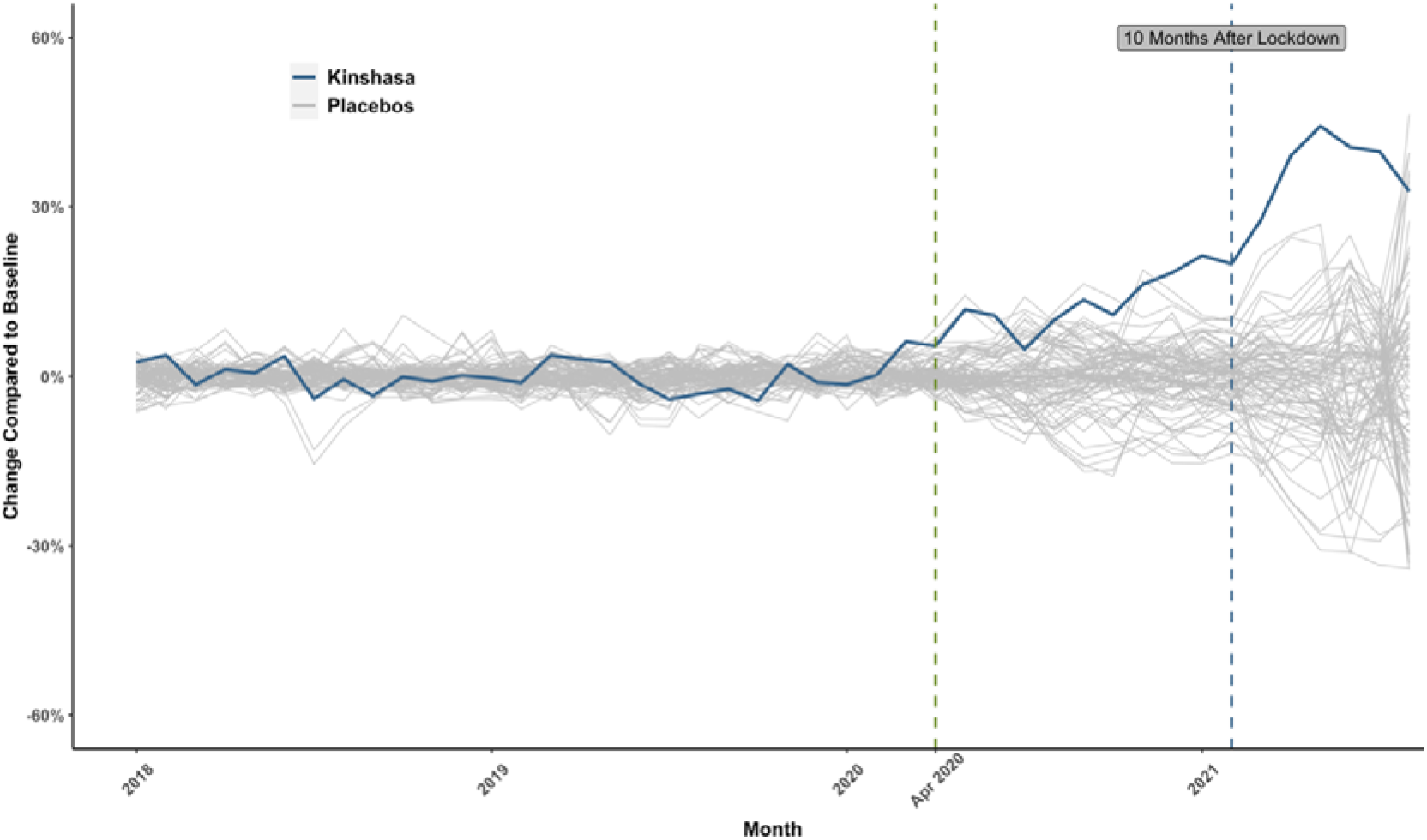
Placebo Test.

## 4. Discussion

This study investigated the impact of lockdown measures, above and beyond other control measures implemented in the DRC, on fertility, which we proxy by the number of births in health facilities. We find that starting approximately 7 months after the lockdown, and lasting for another 5 months, there were very large increases in the number of women giving birth in Kinshasa. Specifically, we observed approximately 5,000 additional births per month, a nearly 45% increase in deliveries relative to baseline. We also observe important increases in complimentary services that are normally associated with births, especially receipt of PNC within 6 hours of delivery and doses of the BCG vaccine. We did not see any significant increases in ANC visits, nor did we see any major increases in visits for diarrhea or total outpatient visits. Our placebo test provides very strong corroborating evidence that the effects observed in Kinshasa were distinct and more extreme than those observed in the rest of the country. As such, we conclude that the lockdown implemented in Kinshasa in 2020 led to a large increase in fertility. We observed important increases in births for both women over the age of 20 as well as those below, however, proportionally increases were larger for older women than for the younger women.

Given the limitations of our data, we are unable to speculate as to whether the increases in pregnancies we observed were intended or unintended. There is, however, growing evidence globally as well as from the DRC that lockdowns implemented during the COVID-19 pandemic had been associated with large increases in intimate partner violence. A national online survey conducted in August-September 2020 reported that the prevalence of IPV in the DRC was about 11% in the past month (Ditekemena et al., 2021a). The study also found that pregnant women were more likely than other women to report IPV, which is suggestive that the rise of these types of behaviours may have contributed to increases in pregnancies in the DRC.

As previously mentioned, while our identification strategy specifically investigated the effects of lockdowns on fertility, it is important to note that at the time when the lockdown had been implemented in Kinshasa, schools had already been closed both in the capital city and in the rest of the country. As such, our results should be seen as the impact of lockdown above and beyond other public health measures but in a context in which young adolescent girls may have been more vulnerable to getting pregnant. A recent study from Kenya found that adolescent girls that completed school before the start of the COVID-19 pandemic were less likely to have fallen pregnant before completing secondary school, less likely to drop out, and were less likely to have reported that their first sexual experience was desired (Zulaika et al., 2022). While the authors attribute this effect to lockdowns, the two groups of girls in this study were also exposed to the pandemic and all its associated response measures, especially prolonged school closures, not just the lockdown. In our study, while we find increases in pregnancies among younger women, it was by no means only school-aged girls that experienced an increase in pregnancies.

Another important caveat to our findings that is worth discussing is that while all of Kinshasa had been under government mandated directives to greatly reduce movement and social contact, including school and business closures, it was mainly the central commune of Gombe that had been put under a very strict lockdown. However, even so, it is easy to imagine how the closure of Gombe could have had far-reaching effects well beyond those just in Gombe. As a central business district, many people would normally work or commute through this area, thus not just people in Gombe were affected by the policy. Women of husbands who normally would have commuted outside of Kinshasa also would have seen their husbands more than normal. Bars, nightclubs, and other normal places of entertainment in urban centres had all been closed, which likely led to high levels of idleness amongst many. We considered independently evaluating the impact of the lockdown on births just in the Gombe commune, however, given that most of the increased births we observed in our sample occurred at a time in which Gombe was no longer under lockdown, and given the location of the main provincial referral hospital in this area, it was impossible to distinguish the births amongst women who lived in Gombe at the time of lockdown and those that had not.

While we believe our study sheds new light on an important mechanism that may have affected fertility during the COVID-19 pandemic, it is not without limitations. First, we do not directly measure the true number of births but instead proxy fertility by the number of women who give birth in health facilities. We speculate that the proportional increase in the number of births observed in health facilities represents a good reflection of the true changes in pregnancy rates in Kinshasa, rather than just a shift in the proportion of pregnant women who give birth in clinics over time. Several factors allow us to draw this inference with reasonable levels of confidence. First, by 2013-14, institutional delivery rates were already nearly universal in Kinshasa and very high in the rest of the country as well (Ministère du Plan et Suivi de la Mise en œuvre de la Révolution de la Modernité et al., 2014). Given the magnitude of the increases in births we observed, it is very likely that the increases that we observe represent actual increases in pregnancies. Second, following a peak in the number of births, we also see a decline in the number of births after 5 months of increases, which again would be consistent with the effects of a short-run measure like lockdown. Another limitation of our study is that our dataset has less complete coverage of health services delivered in private health facilities, which do account for an important proportion of all institutional deliveries, in particular in Kinshasa (Ministère du Plan et Suivi de la Mise en œuvre de la Révolution de la Modernité et al., 2014). It is possible that there may have been important shifts in the mix of births happening in the public vs the private sector and this could also be affecting our results. While an important shift towards the use of private health services has been documented in other contexts during the pandemic and was even part of the public health response in some countries (Williams et al., 2021), had such a shift occurred, they would have most likely happened in the immediate aftermath of the lockdown and not necessarily many months later. Plus, in general, the shifts in the public vs private mix in deliveries that have been observed in other contexts during previous infectious disease outbreaks were mainly in the direction of more private sector deliveries (Blair et al., 2017), not less, and thus it is also not likely the case that this limitation of our dataset that is driving the large increases that we had observed.

The COVID-19 pandemic led to the adoption of an unprecedented number of policies aimed at containing and mitigating the impact of the virus. Lockdowns were widely used, including by many African countries (Haider et al., 2020). Our study provides evidence that the unintended effects of these policies may have effect long after such policies had been lifted. Before the next infectious disease outbreak, global health experts and national policy makers need to better understand the potential unintended effects of these policies on outcomes, such as fertility, and to develop strategies to mitigate these effects, for example by providing outreach or mobile access to family planning services, support to out of school adolescents, and other support.

## Data Availability

Data are available from the Ministry of Public Health

## List of Abbreviations

1. LMICs: Low- and Middle-Income Countries
2. HICs: High-Income Countries
3. IPV: Intimate Partner Violence
4. SSA: Sub-Saharan African
5. DHS: Demographic and Health Survey
6. HMIS: Health Management Information System
7. DHIS2: District Health Information System 2
8. ANC: Antenatal Care Visits
9. PNC: Postnatal Care Visits
10. BCG: Bacillus Calmette-Guérin
11. DTP-HepB-Hib: Diphtheria-Pertussis-Tetanus-Polio-Hepatitis B-Haemophilus Influenzae Type B
12. SCM: Synthetic Control Method
13. ATT: Average Treatment effect on the Treated
14. ASCM: Augmented Synthetic Control Method

## Appendices

**Table A.1.** Estimated ATTs for deliveries before and after the lockdown period

|  | Month | Estimated ATT (% to the Baseline) |
| --- | --- | --- |
| **Pre-Lockdown Periods** | January 2018 | 301.95 (3%) |
|  | February 2018 | 450.72 (4%) |
|  | March 2018 | -178.53 (-1%) |
|  | April 2018 | 149.58 (1%) |
|  | May 2018 | 72.84 (1%) |
|  | June 2018 | 428.07 (4%) |
|  | July 2018 | -479.28 (-4%) |
|  | August 2018 | -67.32 (-1%) |
|  | September 2018 | -413.28 (-3%) |
|  | October 2018 | -15.42 (0%) |
|  | November 2018 | -106.43 (-1%) |
|  | December 2018 | 20.00 (0%) |
|  | January 2019 | -39.15 (0%) |
|  | February 2019 | -139.92 (-1%) |
|  | March 2019 | 449.85 (4%) |
|  | April 2019 | 362.94 (3%) |
|  | May 2019 | 307.60 (3%) |
|  | June 2019 | -163.26 (-1%) |
|  | July 2019 | -489.25 (-4%) |
|  | August 2019 | -378.00 (-3%) |
|  | September 2019 | -278.49 (-2%) |
|  | October 2019 | -516.45 (-4%) |
|  | November 2019 | 256.24 (2%) |
|  | December 2019 | -130.18 (-1%) |
|  | January 2020 | -172.24 (-1%) |
|  | February 2020 | 24.40 (0%) |
| **Onset of COVID-19** | March 2020 | 741.72 (6%) |
| **Post-Lockdown Periods** | April 2020 | 644.21 (5%) |
|  | May 2020 | 1424.37 (12%) |
|  | June 2020 | 1295.76 (11%) |
|  | July 2020 | 570.17 (5%) |
|  | August 2020 | 1195.76 (10%) |
|  | September 2020 | 1631.45 (14%) |
|  | October 2020 | 1313.70 (11%) |
|  | November 2020 | 1960.49 (16%) |
|  | December 2020 | 2209.20 (18%) |
|  | January 2021 | 2573.61 (21%) |
|  | February 2021 | 2399.18 (20%) |
|  | March 2021 | 3340.70 (28%) |
|  | April 2021 | 4703.25 (39%) |
|  | May 2021 | 5334.73 (44%) |
|  | June 2021 | 4887.01 (41%) |
|  | July 2021 | 4793.05 (40%) |
|  | August 2021 | 3945.07 (33%) |

## Footnotes

1 ^1^ http://who.covid19.int

